# Evaluation of Easy-to-Implement Intervention for Menstrual Pain in a Series of N-of-1 Trials: Study Protocol of the Menstrual Pain Intervention Among Students Study (MPIS)

**DOI:** 10.1101/2025.09.20.25336223

**Authors:** Valeria Tisch, Hannah Neumayer, Stefan Konigorski

## Abstract

**Background:** Dysmenorrhea, or menstrual pain, is a prevalent issue among female university students that negatively influences their productivity, academic performance, and quality of life. Because the conventional treatments, such as non-steroidal anti-inflammatory drugs and hormonal contraceptives, often lead to undesirable side effects, there is a need for alternative, non-pharmaceutical methods. Abdominal self-massage has been shown potential to decrease menstrual cramping, but the research on its efficacy remains limited. Individuals respond differently to physiotherapy interventions. Therefore, we aim to assess the effect of performing an abdominal self-massage regularly on the self-reported perception of menstrual pain and related symptoms of female university students in Germany, with a series of N-of-1 trials.

**Methods:** A non-randomized, unblinded N-of-1 trial is carried out over 60 days among participants who regularly experience dysmenorrhea. Participants start at different times of their menstrual cycle in a control phase (A) followed by an intervention phase (B) and a possible second control phase (A) depending on the individual cycle length and study start. Daily symptoms are recorded via the StudyU smartphone application. A baseline survey collects demographic, lifestyle and menstrual history information to identify potential effect modifiers. Bayesian multi-level modeling will be used to estimate the intervention’s effects on the individual and population level. Free text descriptions will be analyzed exploratively with a large language model-based natural language processing pipeline.

**Discussion:** This study investigates abdominal massage as a potential self-care practice for managing menstrual symptoms. The results may benefit the individual well-being and contribute to the exploration of a more holistic approach to menstrual health.

**Trial Registration:** ClinicalTrials.gov NCT07155291

## 1. Background

Dysmenorrhea, more commonly referred to as painful menstruation or menstrual cramps, is a condition characterized by intense cramps in the lower abdomen and is the most common menstrual complaint among women [1]. It is often accompanied by other symptoms as well, such as nausea, bloating, fatigue, or headaches. Dysmenorrhea can be categorized into two types: primary dysmenorrhea, which occurs without a known underlying disease, and secondary dysmenorrhea, which occurs due to a pelvic pathology like endometriosis [2]. The prevalence of dysmenorrhea reported in the research varies significantly, ranging from 16% to 91%. Severe cases are less common, affecting 2% to 29% depending on the definition of severity. [3] The highest prevalence is seen in young females of 17–24 years, with estimates between 67% and 90% [4], [5]. Even though the literature reports substantially different numbers, all of them underline dysmenorrhea as a frequent issue. In particular, severe dysmenorrhea has been shown to influence health-related quality of life and work productivity [3]. The overall life satisfaction is rated poorer during menstruation by affected women [6]. In students, dysmenorrhea negatively impacts their academic performance, which manifests itself in a decreased classroom participation and ability to focus [5]. Furthermore, students attend university less often and participate less in sports and social activities [7].

Approved treatments include Non-Steroidal Anti-Inflammatory Drugs (NSAIDs) as the main option for pain relief and the use of combined oral contraceptives [8]. However, these treatments can cause a number of undesirable short-term and long-term side effects, such as gastrointestinal issues like gastritis, nausea, and diarrhea, or neurological problems, including migraines, depression, and fatigue [9]. Alternative options with less or no adverse effects should be explored. Especially, physical therapy has proven to be effective. Given its overall health benefits and minimal risk of side effects, physiotherapy can be used alone or in combination with other methods. It usually improves blood circulation and reduces muscle tension. [10] One possible easy-to-implement physical therapy intervention is abdominal massage that can be performed by patients themselves [11]. Previously, it has been shown to alleviate symptoms like abdominal and back pain, as well as fatigue [12].

However, research in this respect remains sparse and faces some challenges. The possible response within a population to this type of intervention can vary greatly. Additionally, there are methodological barriers that often prevent the full integration of women’s individual biological differences (different cycle lengths, irregular menstruation) into standardized clinical studies [13]. Accordingly, exploring study designs that accommodate these complexities is necessary. Given the variability in cycle lengths and menstruation experiences, the rigid structure of traditional randomized controlled trials (RCTs) can be impractical. In contrast, N-of-1 trials are particularly well-suited in case of such heterogeneity [14]. These trials compare an intervention’s effect against control phases within a single patient. This within-subject design allows to investigate personalized treatment protocols and allows inference both on individual-level treatment effects [15] and population-level where it requires fewer participants for inference on aggregated average effects compared to standard RCTs [16]. Furthermore, many women already have strong preferences for how to manage menstrual symptoms, or may develop such preferences throughout a study. This makes forced randomized allocation ethically questionable, because it may withhold treatments that participants found helpful and does not allow them to integrate their prior experience into their decision-making about whether to follow a treatment and a specified phase order. [17] Finally, study participants typically do not obtain any personalized feedback on their study results in standard RCTs, but can get to know which treatment worked best for them in N-of-1 Trials. Therefore, we apply a non-randomized, unblinded, experimental N-of-1 trial design in our study. The data gathered at the end of the trials allows for a detailed statistical analysis. On the individual level, health outcomes collected in the intervention phases can be compared to those in the control phases. To assess the outcomes at the population level and make inference on average effects, the series of N-of-1 trials of multiple participants can be aggregated.

In the following, we present the study protocol for conducting a series of N-of-1 trials to investigate the effects of an abdominal self-massage routine on the perception of menstrual pain among female university students residing in Germany, estimating both individual-level and population-level treatment effects. Also, we will analyze participants’ free-text answers utilizing a large language model (LLM)-based natural language processing (NLP) approach to detect additional symptoms and the sentiment of their descriptions, how it changes over time, and compare to the structured measures.

### 1.1. Objectives

The primary goal of this study is to explore whether performing an abdominal self-massage on a regular basis can reduce menstrual pain. First, we will focus on analyzing individual-level effects. The impact of the intervention will be compared to its absence in terms of menstrual pain (primary outcome), medication intake reduction, improvement of related symptoms and mood (secondary outcomes) within a series of N-of-1 trials. Second, we will also estimate effects at the population level for the same outcomes. We will incorporate factors that may modify the impact of the intervention, such as body mass index (BMI), general health status, belief in massage effectiveness and more (potential effect modifiers). Finally, we will examine free-text responses written by participants qualitatively with LLM-based NLP methods. As abdominal self-massage is not directly applied but provided through the StudyU app, participants may choose to not perform the massage so that this study investigates the effectiveness under real-world conditions.

## 2. Methods

### 2.1. Study Design

To assess the effect of providing abdominal self-massage compared to providing no intervention on perceived menstrual pain, a series of N-of-1 trials is carried out. A study design diagram is provided in Figure 1. The recruitment process spans over one month. Each trial consists of a screening for eligibility and a baseline survey, followed by a 60-day N-of-1 trial. The Standard Protocol Items: Recommendations for Interventional Trials (SPIRIT) checklist was used for study protocol development [15]. The findings are reported according to the [15] guidelines for N-of-1 trials [15]. The study was approved by the ethics committee of the University of Potsdam (UP) (Approval No. 8/2025).

**Figure 1.**
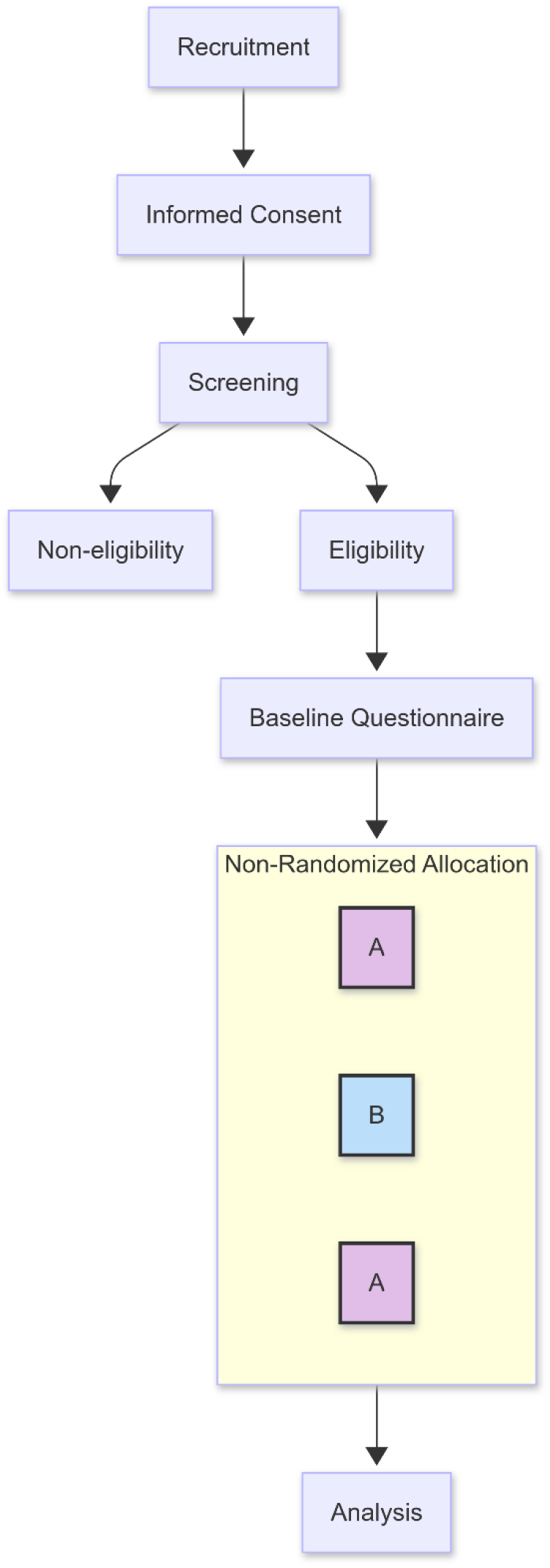
Overview of participant flow and study design.

### 2.2. Intervention

All participants are encouraged to follow the same treatment protocol during the intervention phase (Phase B) daily. The treatment consists of providing an instructional video for self-massage through the StudyU app, in more detail a visceral abdominal self-massage with circular motions and applying pressure to certain points, performed for at least 5 and up to 20 minutes. Participants can determine the duration based on their preference. Previous research has shown that 5 minutes may already be sufficient to achieve results, while a longer duration could enhance effects, but should remain feasible for participants [18]. The StudyU app provides an instructional video as well as a written step-by-step guide with drawings on the proper technique. The massage technique and video were developed by a certified physiotherapist.

During the control phase (Phase A), participants log their symptoms but are not provided with the instructional video for self-massage and are not encouraged to perform the massage.

### 2.3. Participants

The study population consists of female university students residing in Germany who regularly experience menstrual pain (dysmenorrhea) and may also report other related symptoms (e.g., nausea, fatigue, and headaches). Participants are recruited via email through university mailing lists and at German universities. The recruitment period spans one month. Interested individuals can access the study information and necessary consent forms via a universal link. A study landing page was developed for that (https://mpistudy.de/). If participants are interested, they may click a participation link where they can find the necessary study information together with a consent form. After providing their informed consent, participants undergo a screening process, and if they are eligible, they are forwarded to the baseline survey. Upon completing, participants can begin the study immediately. If they have any questions, they may reach out to the principal investigator via email or telephone.

#### 2.3.1. Inclusion Criteria

- Female university student living in Germany
- Having consistently experienced menstrual pain and other symptoms primarily during menstruation for at least the last three cycles
- Regular access to a smartphone on which the StudyU application [19] can be installed (https://studyu.health)
- Providing informed consent
- Proficiency in English or German

#### 2.3.2. Exclusion and Termination Criteria

- Age under 18 years or over 45 years
- Participation in a different intervention study during the duration of this study
- Use of hormonal treatments affecting the menstrual cycle
- Confirmed or suspected pregnancy
- Presence of contra-indicative disorders or diseases for massage (e.g., active inflammatory bowel disease)
- Severe psychiatric conditions impairing informed consent or reliable participation
- Substance abuse (e.g., alcohol, drugs)
- Planned surgery within the next four to five months
- Doctor’s advice against proceeding with the intervention

### 2.4. Procedure

The study collects anonymized self-reported data and is carried out entirely online. Figure 1 outlines the study design and the participants’ flow throughout the trials. Participants are recruited through email lists of German universities and students’ boards for one month. The three steps: informed consent, screening, and baseline survey, shown in Figure 1, are all conducted over UP Survey (https://survey.uni-potsdam.de/), which the participants can access via the universal landing page link at https://mpistudy.de/. First, interested individuals must confirm their acknowledgment of the study information and provide informed consent. If they wish to receive the information sheet and consent forms via email, they must also provide separate consent for the processing of their email address through a different site to ensure their anonymity. To verify interested individuals’ eligibility, the participants must complete a screening process based on the predefined inclusion and exclusion criteria. Those who qualify are forwarded to the baseline survey. This survey records the female students’ characteristics to collect demographic data and explore potential effect modifiers.

Upon completing the baseline survey, participants receive a random, unique invitation/linkage code on the final screen, along with a link to a contact form where they can request corrections or data deletion. In this contact form, they are asked to provide their invitation code but are not required to disclose any identifying information, such as email addresses. This invitation code enables participants to enroll in the Menstrual Pain Intervention Among Students (MPIS) study via the StudyU application and allows researchers to connect the baseline data collected over UP Survey with the app-recorded data. Participants also receive instructions on how to download and install the StudyU app, which is freely available on the App Store and Google Play Store.

All participants follow an AB(A) intervention sequence, where A represents the control phase and B the intervention phase. Participants receive daily push notifications to encourage tracking and adherence. Since menstrual cycle lengths may vary among participants and fluctuate even within the same individual, the exact timing of each phase is unpredictable. To ensure that each participant tracks at least two, potentially three, menstruations during the study period, the study is designed to last 60 days. While all cycle days are analyzed, the menstruation phase is of our primary interest. Participants begin with a control period (A) during their follicular phase (before ovulation), ovulation, or luteal phase (after ovulation but before menstruation). The important condition is that they start while not menstruating.

The duration of the phases depends on the participant’s cycle length and the timing of their enrollment. Figure 2 illustrates two example scenarios with an identical cycle length but different intervention and control phase durations due to enrollments at different times. The study phases adjust to the participant’s cycles, accommodating various cycle lengths. However, we recommend that participants start only a couple of days before their next menstruation, if they can predict it, to ensure they track at least two menstruations within the 60 days in case someone has a longer cycle length (>30 days). To detect the current phase assignment of a participant, they must indicate which stage of their menstrual cycle they are currently in relative to the study timeline. Only if the participant indicates that their first menstruation has ended but their second has not, the app will display the intervention tutorial. Once the minimum required number of Participant’s Reported Outcomes (PROs) is achieved and the trial is completed, participants can access an automatically generated report detailing their data collected via the StudyU application.

**Figure 2.**
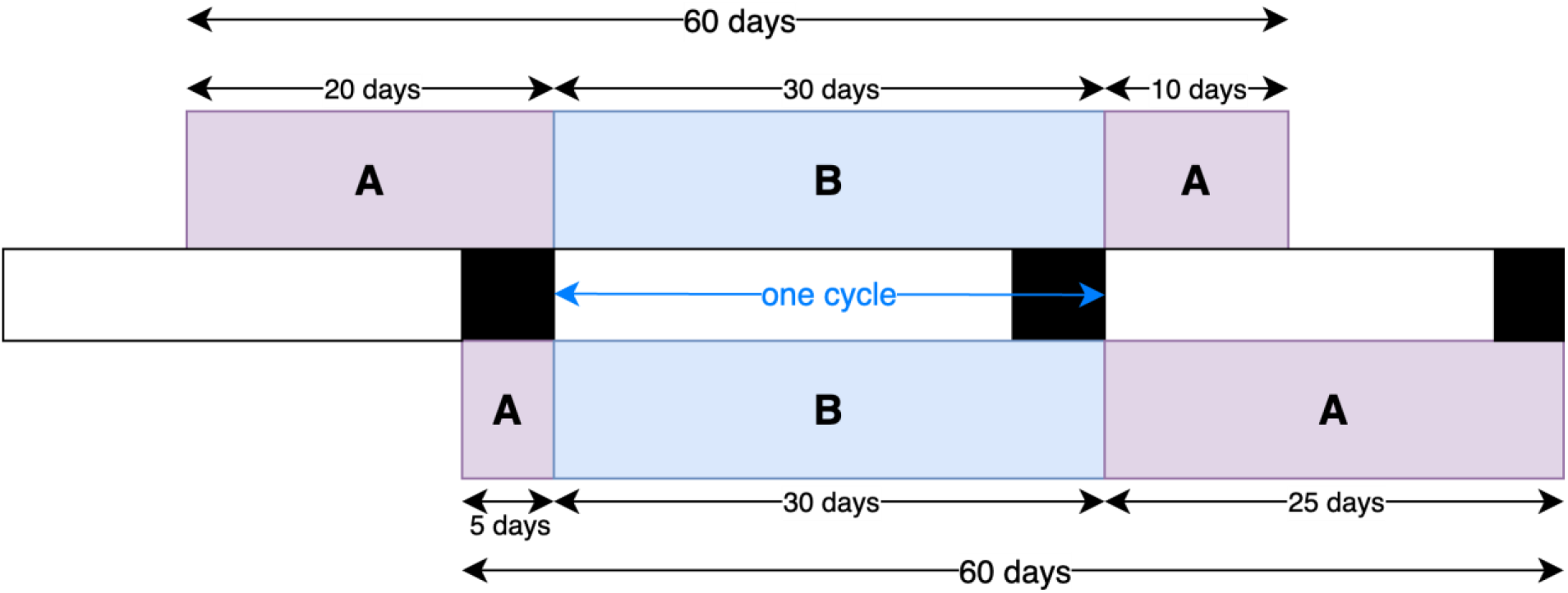
Flexible AB(A) study design with varying phase lengths across participants, showing two example scenarios. Black boxes indicate menstruation, white boxes indicate non-menstruating days. Control phases (A) are purple, intervention phases (B) are blue.

### 2.5. Outcomes

Initial participant data is collected through the baseline survey via the UP Survey web tool. During the N-of-1 trials, the PROs are collected daily after 4 p.m. using the StudyU application. Participants first answer a question whether they had menstrual cramps or other related symptoms on that day. If this question is answered positively on a given day, the other outcome measures are collected, too. Otherwise, the corresponding questions are not displayed. Participants receive daily reminders via mobile push notifications.

#### 2.5.1. Primary Outcome Measure Menstrual Pain Severity

Participants assess the following question on a Visual Analogue Scale (VAS) from 0 to 10 (0=no pain, 10=worst pain imaginable) daily through the StudyU app: *“How severe was your worst menstrual pain today?”*

#### 2.5.2. Secondary Outcome Measures Medication Use

As medication use changes the perception of pain, it is essential to consider this detail when assessing symptom severity as well. Therefore, participants answer the following question daily through the StudyU app: *“How many painkiller tablets did you take today?”* Participants are supposed to indicate the whole number of tablets, regardless of whether they took half a tablet.

##### Dysmenorrhea Symptoms

Since dysmenorrhea is typically accompanied by additional symptoms [2], participants need to indicate if they have experienced any of the following, daily through the StudyU app:

- Cramps
- Lower back pain
- Nausea, vomiting, or diarrhea
- Painful urination
- Painful bowel defecation
- Bloated belly
- Painful intercourse
- Fatigue or brain fog
- Fainting
- Headaches

##### Mood Impact

Furthermore, participants are requested to rate how the symptoms affect their mood on a numeric 5-point Likert-Scale ranging from 0 to 4 (0=*no impact at all*, 4=very strongly negatively), daily through the StudyU app: *“How did your symptoms affect your mood today?”*

#### 2.5.3. Exploratory Outcome Measures Qualitative Diary Entry

Lastly, participants have the option to respond daily through the StudyU app with a free-text diary entry limited to 1000 characters, which will serve for the LLM based NLP analysis: *“How are you feeling today? Please describe your symptoms in your own words. Feel free to include any details about your physical and emotional well-being.”*

#### 2.5.4. Effect Modifiers and Additional Variables in Baseline Survey

Additional data is collected through the baseline self-reported survey to assess female students’ characteristics and investigate subgroups and potential effect modifiers that may influence the study outcomes. Variables include:

- Age group: Participants are categorized into age groups to examine the relationship between age and the risk of dysmenorrhea, previously reported as inverse [20], [3]. The age groups (18-25, 26-32, 33-39, 40–45 years) generally reflect biological transitions and changes in life stages.
- Occupation: Participants indicate whether they work alongside studying, as occupational stress may influence the severity of dysmenorrhea and intervention adherence [21], [22].
- BMI: Height and weight are recorded to calculate BMI=weight in kg/(height in m)^2^, as there may be a link between BMI and increased severity of dysmenorrhea [23],[24].
- General Health Status: Rated on a 5-point Likert scale from 5 (Excellent) to 1 (Poor), this measure evaluates participants’ overall health, which may affect menstrual pain and response to the intervention.
- Lifestyle variables:
  ▸ Physical activity: Assessed using selected items adapted from the Global Physical Activity Questionnaire (GPAQ) [25], participants report the typical frequency and duration of vigorous and moderate activities both outside of and during menstruation. Physical activity has been shown to alleviate symptoms of dysmenorrhea [26]. However, the relationship is likely complex, as activity intensity and timing may matter.
  ▸ Alcohol consumption: Evaluated with the first question of the Alcohol Use Disorders Identification Test, Question 1 (AUDIT-Q1) [27]: *“How often do you have a drink containing alcohol?”* Alcohol consumption may influence factors that affect menstrual symptoms, like pain perception and sleep quality. [28]
  ▸ Smoking status: Recorded as yes, no, or rarely. Given its inflammatory effects, smoking may present a risk factor for dysmenorrhea [3].
  ▸ Massage experience and expectation: Participants document their prior experience with abdominal massage, belief in its effectiveness, and interest in performing the intervention. These measures are relevant for adherence and placebo effects.
  ▸ Menstrual history: Details include age at menarche, age at pain onset, cycle length, typical level of menstrual pain on a VAS, parity, and any previous suspicions or confirmed diagnoses of endometriosis or adenomyosis. Earlier menarche, nulliparity, and long cycles have been associated with a higher risk of dysmenorrhea [3],[28]. Endometriosis and adenomyosis represent secondary causes of dysmenorrhea [2].

### 2.6. Data Management

Data collection and management adhere to the standards set by the European General Data Protection Regulation (GDPR). A randomly generated linkage code is issued automatically at the end of the baseline survey. It allows participants to log into the Menstrual Pain Intervention Among Students (MPIS) study within the StudyU application. It allows researchers to connect data from the UP Survey tool and StudyU questionnaires. Through the two-step setup of participants signing up for the study and then receiving the link to the baseline survey and all further steps, no person-identifying data is collected that can be linked to the other data collected throughout the study. This has also been considered in how participants can request a correction or deletion of their data through a contact form, where they only need to provide their linkage code.

Participants are required to install the StudyU application on their smartphones to complete daily questionnaires. The app, developed and maintained by the Hasso-Plattner-Institute (HPI) in Potsdam, sends reminder notifications. Before participation, individuals must consent to the app’s terms of use, which outline data storage and publication policies. The StudyU application is designed to avoid collecting identifiable information and gather only the data necessary for the study. Participants are informed that they can uninstall the app after the study ends.

Data collected over UP Survey is stored securely on a server at the UP. Data from the StudyU application is stored securely on a backend managed by the HPI. We plan to make anonymized data publicly available, either fully or partially, in a recognized research data repository to support further scientific research. Data obtained through the StudyU application will be published in its designated publicly available repository (https://designer.studyu.health). In compliance with good scientific practice, published data must remain traceable to the original data for a minimum of 10 years. Any unpublished data will be permanently deleted from both the UP and HPI servers six months after study completion.

### 2.7. Estimand

The question to assess intensity of dysmenorrhea and all other questions are provided daily between 4 p.m. local time until the end of the day on the StudyU app. On intervention days, participants are asked to complete the assessment after performing the intervention. To support adherence during the intervention phase, participants receive daily push notifications at 10 a.m. However, they can choose when to perform the intervention exactly and can also opt to not perform the intervention. Actual execution of the intervention is not directly monitored, meaning actual execution data is unavailable. As a result, this study evaluates the immediate effect of providing the intervention, i.e. the effect of access to intervention instructions, on the health outcomes. Our primary interest lies in comparing the effect of regularly implementing the intervention versus not implementing it at all. Under the assumption of full adherence, the effects of receiving the intervention instructions are considered equivalent to the effects of the intervention itself.

We further assume that the treatment effect is largely stable over time, with negligible carry-over effects into the next cycle. While time-varying factors, such as seasonal changes and the academic calendar, are acknowledged, they are not expected to significantly influence the outcomes. Thus, they will not be accounted for in the statistical analysis and the sample size calculations. To evaluate effectiveness, both individual and average causal intervention effects across all participants will be estimated and tested.

### 2.8. Sample Size

To determine the required sample size of individuals for inference on the population-level average effect of an intervention on perceived dysmenorrhea, measured on a VAS from 0 to 10, we performed a highly conservative sample size calculation based on a two-sided paired-sample t-test. That is, under the most conservative assumption that daily menstrual pain measurements might not provide high information, they can be averaged to provide one average menstrual pain level under intervention and one average pain level under no intervention for each study participant, which can then be analyzed on the population-level using a paired t-test. We calculated effect sizes from three comparable studies [12], [11], [29] investigating similar interventions for dysmenorrhea, and aggregated these values with weights based on their respective sample sizes. Conservatively, we assume a standardized intervention effect size of Cohen’s d=0.48, which is slightly more than half of the actual aggregated value. We set the significance level α to 0.05 and the desired statistical power to 0.75. Based on these parameters and using the pwr.t.test function in the pwr package in R, the required sample size is 33 participants. Accounting for an anticipated dropout rate of 30%, we aim to recruit 48 participants. Note that in the actual population-level and individual-level analyses (see below), we will include all daily pain measures, which will lead to much more efficient treatment effect estimates. For individual-level inference, in a naïve analysis, 60 data points would allow to estimate a 95% confidence interval around a clinically meaningful decrease of 0.5 units, assuming a standard deviation of 1 equal across both groups, with equal allocation ratio, with width 0.7, i.e., would allow to detect clinically meaningful effects on the individual level using frequentist analyses. This was computed using the prec_meandiff function in the presize package in R.

### 2.9. Statistical Analysis

Before starting the statistical analysis, a detailed Statistical Analysis Plan (SAP) will be created and published on ClinicalTrials.gov. Statistical analysis will be performed using Python and Bayesian models will be fitted with Markov Chain Monte Carlo (MCMC).

First, descriptive analyses will include descriptive statistics and exploratory plotting of all the distribution of all variables of interest. Next, the primary analysis aims to make inference both on individual-level and population-level effects of abdominal self-massage on dysmenorrhea pain levels within a series of N-of-1 trials, following an intention-to-treat (ITT) approach. On the individual level, the posterior distribution of treatment effects will be estimated using Bayesian linear regression models, fitted separately for each study participant. These estimates should be considered naive as they do not account for trends seen in other participants. In addition, we will fit a Bayesian linear model with autoregressive errors of order 1 (AR1). We will consider an average decrease in menstrual pain by 0.5 points on the 0-10 scale clinically meaningful, and will classify participants as responders to the self-massage treatment if the posterior probability of reaching at least a clinically meaningful effect is at least 70%. On the population level, all individual trial data will be combined in a Bayesian multi-level mixed model to estimate both the average effect of the intervention and the variability between and within participants. Similarly to the individual-level analysis, the posterior distribution of the population-level average treatment effect will be estimated, and an AR1 error structure will be used to account for time dependencies. As effect modifier analysis, we will explore potential effect measure modifiers. Using Bayesian multi-level models, we will explore whether participants’ characteristics, such as baseline worst pain, occupation, and belief in massage effectiveness influence individual responses to the intervention. All available data will be used for each participant, independently of whether data is missing or the participant dropped out of the study. In the AR1 models, such possible data gaps will be considered. We will perform standard model checks and checks of convergence of the MCMC.

Participants’ free-text diary responses will undergo an exploratory NLP analysis. The collected data will be structured to allow further analysis, capturing metadata such as participant IDs, timestamps, and intervention phases. As we expect that the dataset is too small for traditional unsupervised methods such as BERTopic [30], the main analysis of the textual responses will rely on LLMs. The answers will be scored using supervised prompting across different dimensions, like pain intensity and emotional well-being. Beyond that, content extraction will be performed to collect symptom information to see how these are described or if symptoms may be missing on the Dysmenorrhea Symptom List (DSL). The NLP-derived quantitative outcomes will be compared to the structured measurements to assess their alignments. Furthermore, the scored dimensions will be analyzed temporally to identify phase-specific changes and detect shifts in the scored dimensions or symptoms.

### 2.10. Monitoring and Safety

Participation in the study is entirely voluntary, and participants may withdraw at any time without providing a reason. At this point, they can choose to delete all collected data or leave the collected data for analysis. Participants can also drop out of the study by simply not answering any more questions in the StudyU app. Exclusion criteria have been established to minimize potential harm. Participants meeting these criteria at baseline are not enrolled in the study. The intervention is self-administered, short, and of low intensity, and is only to be performed over one cycle. As similar previous studies have reported positive effects and no adverse effects, the risk of adverse outcomes is considered negligible. However, participants are encouraged to report any adverse reactions. The study physician and principal investigator can both be contacted in the case of any concerns or questions.

## 3. Discussion

The primary aim of the MPIS study is to investigate the effects of an abdominal self-massage routine on perceived menstrual pain among female university students in Germany using a series of N-of-1 trials with a non-randomized, unblinded AB(A) design (A: control, B: intervention). Participants will rate their daily dysmenorrhea pain intensity and related symptoms via the StudyU smartphone application. Bayesian analyses will estimate both individual-level and population-level treatment effects and assess potential effect modifiers such as baseline worst pain. Additionally, free-text diary responses will undergo qualitative exploratory analyses using NLP to identify meaningful patterns in symptom descriptions and emotional experiences.

One key strength and innovation of our study design is the use of N-of-1 trials, which enable both individual and group-level evaluation, and are especially important in the study of menstrual pain by their ability to incorporate flexible interventions and study designs. By comparing intervention and control periods within the same participant, this approach effectively addresses the variability in how individuals respond to physiotherapy interventions, especially regarding their menstrual pain perception. Therefore, this study may help participants evaluate whether self-massage works for them. Another strength is that the intervention is guided by a certified physiotherapist through an instructional video. Aggregating data of the series of N-of-1 trials allows for inference on the group-level intervention effect with fewer participants than traditional RCTs, and despite possible differences in individual trial protocols. This is especially important since RCTs often struggle to account for variations in menstrual cycle lengths. A unique methodological strength is the application of NLP techniques to exploratively analyze free text responses, which can capture more nuanced insights than traditional multiple-choice questionnaires. Compared to manual qualitative analysis, this approach reduces bias and inefficiencies. In addition, insights gained from the NLP analysis are compared to the quantitative outcomes. Although small in scale, this study contributes to the historically underrepresented field of women’s health research. It is designed to ethically collect sensitive menstrual health data, with plans for dataset publication to support future research in this field.

Nevertheless, several limitations to the study design must be considered. Because there is no blinding or randomization, participants may be biased toward perceiving an effect and experience a placebo response, which may influence their outcome ratings. Although the study design adapts to individual menstrual cycle variability, it is limited to 60 days. In rare cases, unpredictable and fluctuating cycle lengths may result in menstruation not being captured during the intervention cycle. Moreover, restricting recruitment to German female university students limits generalizability to broader demographics and cultural contexts. Uncontrolled external and possibly time-varying factors such as stress, lifestyle changes, or academic schedules, could influence menstrual symptoms independently of the intervention. Although we capture participants’ general lifestyle through the baseline survey, these factors may change over time and are not continuously measured. Hence, our analyses are all based on the assumptions of no time-varying confounders and of no time-varying treatment effect and will estimate marginal effects.

Despite this, the intervention’s simplicity and self-administered nature promote self-efficacy while minimizing reliance on medical professionals, making it practical for real-world and daily life application. Additionally, each participant will receive their individual N-of-1 trial results. These factors may encourage participant adherence and potential long-term adoption post-study if the intervention proves effective for them. A risk of longitudinal studies is participant dropout, especially for digital studies since participants must complete the study without personal supervision. However, this risk is mitigated by a conservative dropout estimate of 30% to ensure robust statistical power. Furthermore, the fully digital execution of the study via an app, including daily reminders, simplifies participation and data collection.

Dysmenorrhea is highly prevalent among young female adults, affecting life satisfaction, productivity, and academic performance. Conventional treatments may not be suitable for everyone due to their side effects. This study explores abdominal self-massage as a potential alternative for improving menstrual symptoms. If demonstrated to be effective, it could enhance individual well-being while contributing to a more holistic approach to menstrual health management.

## Data Availability

After the completion of the study, anonymized data will be available through a publicly accessible research data repository within the StudyU application.

## Declarations

## Acknowledgements

The authors would like to thank the physiotherapist Katherina Bansemer for developing and recording the intervention tutorial; Johannes Vedder for implementing the necessary new features in the StudyU app; Valentin Vetter for providing materials that served as a reference for our study; Dr. Philipp Stoffers for overseeing our study as a study physician.

## Trial Status

The trial has not yet started. Recruitment is planned to begin end of September 2025.

## Ethics Approval and Consent to Participate

All participants provided informed consent, and the study was approved by the Ethics Committee of the University of Potsdam (Approval No. 8/2025).

## Access to Data

After the study’s completion, anonymized data will be available through a publicly accessible research data repository within the StudyU application (see https://designer.studyu.health).

## Authors Contribution

VT: Initial idea, conceptualization and study design, intervention development, development and implementation of questionnaires, methodology and statistical planning, statistical analysis, trial registration and data management plan, writing the initial draft, review and editing, visualizations (figures and tables), final approval for publication.

HN: Revision of intervention and questionnaires, methodology and statistical planning, review and editing, supervision and oversight, final approval for publication.

SK: Initial idea, conceptualization and study design, methodology and statistical planning, review and editing, supervision and oversight, final approval for publication.

## Funding

Not applicable.

## Conflict of Interest

The authors declare no conflict of interest.

## Notes

### Competing Interest Statement

The authors have declared no competing interest.

### Clinical Trial

NCT07155291

### Funding Statement

This study did not receive any funding

### Author Declarations

The Ethics committee of the University of Potsdam gave ethical approval for this work (Approval No. 8/2025).

## Bibliography

[1] M. Schoep, T. Nieboer, M. van der Zanden, D. Braat, and A. Nap, “The impact of menstrual symptoms on everyday life: a survey among 42,879 women.,” American journal of obstetrics and gynecology, p. 569, 2019, doi: 10.1016/j.ajog.2019.02.048.

[2] G. Lentz, R. Lobo, D. Gershenson, and V. Katz, Comprehensive Gynecology E-Book. Elsevier Health Sciences, 2012. [Online]. Available: https://books.google.de/books?id=X5KT_w6Nye8C

[3] H. Ju, M. Jones, and G. Mishra, “The Prevalence and Risk Factors of Dysmenorrhea.,” Epidemiologic reviews, vol. 36, pp. 104–113, 2014, doi: 10.1093/epirev/mxt009.

[4] N. Habibi, M. Huang, W. Y. Gan, R. Zulida, and S. morteza Safavi, “Prevalence of Primary Dysmenorrhea and Factors Associated with Its Intensity Among Undergraduate Students: A Cross-Sectional Study.,” Pain management nursing: official journal of the American Society of Pain Management Nurses, pp. 855–861, 2015, doi: 10.1016/j.pmn.2015.07.001.

[5] M. Armour et al., “The Prevalence and Academic Impact of Dysmenorrhea in 21,573 Young Women: A Systematic Review and Meta-Analysis.,” Journal of women’s health, 2019, doi: 10.1089/jwh.2018.7615.

[6] S. Iacovides, I. Avidon, A. Bentley, and F. Baker, “Reduced quality of life when experiencing menstrual pain in women with primary dysmenorrhea,” Acta Obstetricia et Gynecologica Scandinavica, vol. 93, pp. 213–217, 2014, doi: 10.1111/aogs.12287.

[7] M. Al-Jefout et al., “Dysmenorrhea: Prevalence and Impact on Quality of Life among Young Adult Jordanian Females.,” Journal of pediatric and adolescent gynecology, pp. 173–185, 2015, doi: 10.1016/j.jpag.2014.07.005.

[8] H. Zahradnik, A. Hanjalic-Beck, and K. Groth, “Nonsteroidal anti-inflammatory drugs and hormonal contraceptives for pain relief from dysmenorrhea: a review.,” Contraception, pp. 185–196, 2010, doi: 10.1016/j.contraception.2009.09.014.

[9] J. Marjoribanks, R. O. Ayeleke, C. Farquhar, and M. Proctor, “Nonsteroidal anti-inflammatory drugs for dysmenorrhoea.,” The Cochrane database of systematic reviews, vol. 7, p. CD1751, 2015, doi: 10.1002/14651858.CD001751.pub3.

[10] R. López-Liria et al., “Efficacy of Physiotherapy Treatment in Primary Dysmenorrhea: A Systematic Review and Meta-Analysis,” International Journal of Environmental Research and Public Health, vol. 18, 2021, doi: 10.3390/ijerph18157832.

[11] S. Azima, H. R. Bakhshayesh, M. Kaviani, K. Abbasnia, and M. Sayadi, “Comparison of the Effect of Massage Therapy and Isometric Exercises on Primary Dysmenorrhea: A Randomized Controlled Clinical Trial,” Journal of Pediatric and Adolescent Gynecology, vol. 28, no. 6, pp. 486–491, 2015, doi: 10.1016/j.jpag.2015.02.003.

[12] N. Ozturk, E. G. Öter, and M. K. Eken, “The effect of abdominal massage and stretching exercise on pain and dysmenorrhea symptoms in female university students: A single-blind randomized-controlled clinical trial,” Health Care for Women International, vol. 44, pp. 621–638, 2022, doi: 10.1080/07399332.2022.2061973.

[13] S. Geller, A. Koch, P. Roesch, A. Filut, E. Hallgren, and M. Carnes, “The More Things Change, the More They Stay the Same: A Study to Evaluate Compliance With Inclusion and Assessment of Women and Minorities in Randomized Controlled Trials,” Academic Medicine, vol. 93, p. 1, 2017, doi: 10.1097/ACM.0000000000002027.

[14] J. Nikles and G. K. Mitchell, “The Essential Guide to N-of-1 Trials in Health,” in Springer Netherlands, 2015. [Online]. Available: https://api.semanticscholar.org/CorpusID:33597874

[15] S. Vohra et al., “CONSORT extension for reporting N-of-1 trials (CENT) 2015 statement,” BMJ (Clinical Research Ed.), vol. 350, p. h1738, 2015, doi: 10.1136/bmj.h1738.

[16] S. Punja et al., “N-of-1 trials can be aggregated to generate group mean treatment effects: a systematic review and meta-analysis.,” Journal of clinical epidemiology, vol. 76, pp. 65–75, 2016, doi: 10.1016/j.jclinepi.2016.03.026.

[17] R. L. Kravitz and N. Duan, “ Conduct and Implementation of Personalized Trials in Research and Practice,” Harvard Data Science Review, vol. 0, no. Special Issue 3, Sep. 2022.

[18] T. Marzouk, A. Elnemer, and H. Baraka, “The Effect of Aromatherapy Abdominal Massage on Alleviating Menstrual Pain in Nursing Students: A Prospective Randomized Cross-Over Study,” Evidence-based complementary and alternative medicine: eCAM, vol. 2013, p. 742421, 2013, doi: 10.1155/2013/742421.

[19] S. Konigorski et al., “StudyU: A Platform for Designing and Conducting Innovative Digital N-of-1 Trials,” 2023, doi: 10.25932/publishup-58037.

[20] A. Weissman, A. Hartz, M. Hansen, and S. Johnson, “The natural history of primary dysmenorrhoea: A longitudinal study,” BJOG: an international journal of obstetrics and gynaecology, vol. 111, pp. 345–352, 2004, doi: 10.1111/j.1471-0528.2004.00090.x.

[21] D. C. Christiani, T. Niu, and X. Xu, “Occupational Stress and Dysmenorrhea in Women Working in Cotton Textile Mills,” International journal of occupational and environmental health, pp. 9–15, 1995, doi: 10.1179/OEH.1995.1.1.9.

[22] M. Kordi and M. T. Shakeri, “The Relationship between Occupational Stress and Dysmenorrhea in Midwives Employed at Public and Private Hospitals and Health Care Centers in Iran (Mashhad) in the Years 2010 and 2011,” Iranian journal of nursing and midwifery research, vol. 18, pp. 316–322, 2013.

[23] S. D. Harlow and M. Park, “A longitudinal study of risk factors for the occurrence, duration and severity of menstrual cramps in a cohort of college women,” British Journal of Obstetrics and Gynaecology, vol. 103, no. 11, pp. 1134–1142, 1996, doi: 10.1111/j.1471-0528.1996.tb09597.x.

[24] K. T. Zondervan, C. M. Becker, and S. A. Missmer, “Endometriosis,” The New England Journal of Medicine, vol. 382, no. 13, pp. 1244–1256, Mar. 2020, doi: 10.1056/NEJMra1810764.

[25] T. Armstrong and F. Bull, “Development of the World Health Organization Global Physical Activity Questionnaire (GPAQ),” Journal of Public Health, vol. 14, pp. 66–70, 2006, doi: 10.1007/s10389-006-0024-x.

[26] E. Fernández-Martínez, M. Onieva-Zafra, and M. Parra-Fernández, “The Impact of Dysmenorrhea on Quality of Life among Spanish Female University Students,” International Journal of Environmental Research and Public Health, vol. 16, 2019, doi: 10.3390/ijerph16050713.

[27] D. F. Reinert and J. P. Allen, “The Alcohol Use Disorders Identification Test (AUDIT): a review of recent research.,” Alcoholism, clinical and experimental research, pp. 272–279, 2002, doi: 10.1111/J.1530-0277.2002.TB02534.X.

[28] L. Wang et al., “Prevalence and Risk Factors of Primary Dysmenorrhea in Students: A Meta-Analysis,” Value in Health, vol. 25, p., 2022, doi: 10.1016/j.jval.2022.03.023.

[29] N. Sholihah and I. Azizah, “The Effect of Effieurage Massage on Primary Dysmenorrhea in Female Adolescent Students,” Jurnal Info Kesehatan, vol. 18, pp. 9–17, 2020, doi: 10.31965/infokes.Vol18.Iss1.310.

[30] M. Grootendorst, “BERTopic: Neural topic modeling with a class-based TF-IDF procedure,” arXiv preprint 2203.05794, 2022.

